# Exploring the Behavioral Determinants of COVID-19 Vaccine Acceptance among an Urban Population in Bangladesh: Implications for Behavior Change Interventions

**DOI:** 10.1101/2021.04.23.21255974

**Authors:** Md Abul Kalam, Thomas P. Davis, Shahanaj Shano, Nasir Uddin, Md. Ariful Islam, Robert Kanwagi, Ariful Islam, Mohammad Mahmudul Hassan, Heidi J. Larson

## Abstract

**Background:** While vaccines ensure individual protection against COVID-19 infection, delay in receipt or refusal of vaccines will have both individual and community impacts. The behavioral factors of vaccine hesitancy or refusal are a crucial dimension that need understanding to implicate appropriate interventions. The aim of this study was to assess the behavioral determinants of COVID-19 vaccine acceptance and to provide recommendations to increase the uptake of COVID-19 vaccines in Bangladesh.

**Methods:** We employed a Barrier Analysis (BA) approach to examine twelve potential behavioral determinants (drawn from the Health Belief Model and Theory of Reasoned Action [TRA]) of intended vaccine acceptance. We conducted 45 interviews with those who intended to take the vaccine (Acceptors) and another 45 interviews with those who did not have that intention (Non-acceptors). We performed data analysis to find statistically significant differences and to identify which beliefs were most highly associated with acceptance and non-acceptance with COVID-19 vaccines.

**Results:** COVID-19 vaccine Acceptors in Dhaka were different from Non-acceptors in terms of many of their beliefs and responses. The behavioral determinants associated with the behavior included perceived social norms, perceived safety of COVID-19 vaccines and trust in them, perceived risk/susceptibility, perceived self-efficacy, perceived positive and negative consequences, perceived action efficacy, perceived severity of COVID-19, access, and perceived divine will. In line with the Health Belief Model, beliefs about the disease itself were highly correlated with vaccine acceptance, although not the only determinant. Other responses of Acceptors provide clues such as providing vaccination through government health facilities, schools, and kiosks, and having vaccinators maintain proper COVID-19 health and safety protocols as to ways to make it easier to boost acceptance.

**Conclusion:** An effective behavior change strategy for COVID-19 vaccines uptake will need to address multiple beliefs and behavioral determinants, reducing barriers and leveraging enablers identified in this study. The national plans on COVID-19 vaccination should adopt culturally and community label acceptable and appropriate evidence-based behavior change interventions strategies to promote high vaccination coverage and acceptance in all societal structures across the country.

## Introduction

The severe acute respiratory syndrome coronavirus 2 (SARS-CoV-2) pandemic, popularly known as COVID-19, has infected more than 14 million people with more 3 million deaths in 224 countries (1). The pandemic poses an immense threat to the global public health system and is causing social and cultural disruptions (2,3). Bangladesh reported its first clinically-confirmed COVID-19 case on March 8, 2020, and as of 19 April 2021, Bangladesh has reported 718,950 COVID-19 cases with 10,385 deaths (4) and the counts are increasing with the passage of time. The social and economic costs of COVID-19 have been significant in the developing countries like Bangladesh (5). As the pandemic is expected to continue to impose enormous burdens of morbidity and mortality, and to severely disrupt societies and economies, the introduction of effective COVID-19 vaccines is the only clinical preventive measure (6). As of 22 April 2021, Bangladesh has administered 7 080 699 doses of Oxford/AstraZeneca (COVISHIELD) vaccine were administered in the whole country, out of which, 5 714 090 population received their 1st dose, and 1 366 609 competed for their two doses schedule (7). However, like many other countries, the government has focused the vaccination administration to very specific group of people and general public expected to include in the vaccination program subsequently.

Nevertheless, vaccine hesitancy or refusal around COVID-19 vaccines is a growing concern worldwide. The World Health Organization (WHO) identified vaccine hesitancy as one of the top ten global health threats in 2019 (8) and this is not exceptional in the case of COVID-19 vaccines. In Bangladesh, a survey reported that 81% of urban (metropolitan, district and municipalities) people would get vaccinated when a COVID-19 vaccine is available (9). In a multi-country survey, it was found that only 71.5% of participants reported that they would be very or somewhat likely to take a COVID-19 vaccine (10). A rapid systematic review of 23 peer-reviewed studies and 103 additional syndicated surveys around COVID-19 vaccine hesitancy in the US and globally showed that perceived risk, concerns over vaccine safety and effectiveness, doctors’ recommendations, and inoculation history were common factors influencing hesitancy around vaccination (11). Covid-19 vaccine hesitancy was found to be growing between March and November 2020 (12), but improvements in vaccine acceptance have been noted in some parts of the world since November (13). A growing number of studies have identified demographic, socioeconomic, and behavioral factors that are linked with levels of vaccine acceptance (10,14,15). These factors include age and marital status (16,17), level of education and ethnic origin (17–19), previous vaccination with the influenza vaccine (15,20), and gender (16). Moreover, trust, misconceptions, misinformation, and lack of knowledge among the community on vaccine-preventable diseases have been considered influential determinants of the lower level of acceptance (10,11,14,15,21). These factors have influenced vaccine uptake during previous pandemics and outbreaks caused by H1N1, MERS, SARS, and Ebola virus (22–25).

A meta-analysis demonstrated that the use of social psychological health behavioral change models (e.g. the HBM and Theory of Planned Behavior) would be useful for identifying the influencing determinants of vaccine acceptance (21). The use of economic models when studying vaccine acceptance or hesitancy exhibit some shortcomings in describing the determinants (21,26,27). Behavioral studies have shown that the decision to vaccinate is often based on perceived benefits, effectiveness, and perceived risk of vaccine side-effects versus infection (28). Systematic reviews on behavioral determinants of health have shown that the HBM was useful in identifying determinants associated with the acceptance of Human Papillomavirus (HPV) (26) and influenza vaccination uptake (29). This model has also been found to be effective in predicting intention to vaccinate against influenza among health care workers in Jordan (30). Similarly, a study using Theory of Planned Behavior (TPB) (which was developed from the TRA) showed that vaccine intentions were determined by attitudes, subjective norms, and perceived behavioral control regarding vaccinations among the college students (31). In a comparative study of TPB and TRA, it was found that attitudes and perceptions of social support were determinants for HPV vaccination uptake (27).

Vaccine hesitancy and acceptance are complex in nature, and vaccine decisions can vary according to context, time and place (32). Global studies on demographic determinants can have limited value when looking at determinants of COVID-19 vaccine acceptance in a given country, time, and geographical area. The Technical Advisory Group on Behavioral Insights and Sciences for Health of WHO has identified a number of behavioral drivers including enabling environment, social influences and motivation and recommended to contextualize these drivers into national plans of COVID-19 vaccination (33). Understanding how different behavioral attributes affect individual preferences about vaccination at as granular level as feasible can help inform public health authorities about the actionable activities and messages that will be necessary to achieve broader community uptake of vaccines. Therefore, the primary objective of this study was to explore the behavioral determinants of COVID-19 vaccine acceptance among people of different societal structures in urban communities in Bangladesh. The secondary objective of this study is to provide policy recommendation of culturally acceptable behavioral change intervention points to address these determinants to improve COVID-19 vaccine uptake.

## Materials and methods

### Study site and context

This Barrier Analysis study was conducted in different urban areas of Dhaka – the capital city in Bangladesh – from January 09-15 2021. Dhaka is the residence of more than 10.3 million people – 6.29 percent of total population of the country (34). Between 8 March 2020 and 19 April 2021, there were 736,074 COVID-19 confirmed cases with 10,781 reported death (4). While Dhaka makes up about 6.29% of the total population of Bangladesh, almost 71% of the COVID-19 cases in Bangladesh reported as of 19 April in Dhaka (61% in Dhaka city alone) (4) and which found as one of the largest hotspots of spreading COVID-19 (35).

### Study tool

A Barrier Analysis (BA) study was conducted to better understand the behavioral determinants of COVID-19 vaccine hesitancy in Dhaka. BA is a research tool that was developed in 1990 by Davis (36). Based on HBM and TRA, BA studies explore respondents’ beliefs about a behavior. Sometimes certain beliefs about a behavior (e.g., possible COVID-19 vaccination side effects) are common in a population, but are not necessarily associated with vaccine acceptance. BA is meant to, identify the most likely determinants of that behavior(37). A key feature of BA is that responses from those accepting a behavior (‘Doers’ or ‘Acceptors’) are compared with those who are not (the ‘Non-doers’ or ‘Non-acceptors’) to identify behavioral determinants associated with a particular behavior (e.g., handwashing with soap, getting a vaccine) so that practitioners can develop more effective behavior change messages and activities. BA has been used in more than 40% of low-to-middle-income countries and used extensively by World Vision and other organizations during both the Ebola (38) and COVID-19 pandemics (39). The beliefs and other responses regarding behavioral determinants assessed during BA (see Box 1), are identified with a focus on the most actionable findings. The other details of BA approach can be found elsewhere (40–45). There are BA studies in the peer-reviewed literature on exclusive breastfeeding (46) HWWS among internally displaced women in the Kurdistan region of Iraq (40) timely oral polio vaccination agricultural extension behaviors in India (47), dietary salt reduction in Nepal (48) transition from the lactational amenorrhea method to other modern family planning methods in Bangladesh (36) and cervical cancer screening in Senegal (49).

### Questionnaire development

This study modified the standardized Barrier Analysis questionnaire from the *Designing for Behavior Change* (DBC) training manual (45).

#### Box 1

**Key definitions based on Barrier Analysis approach**

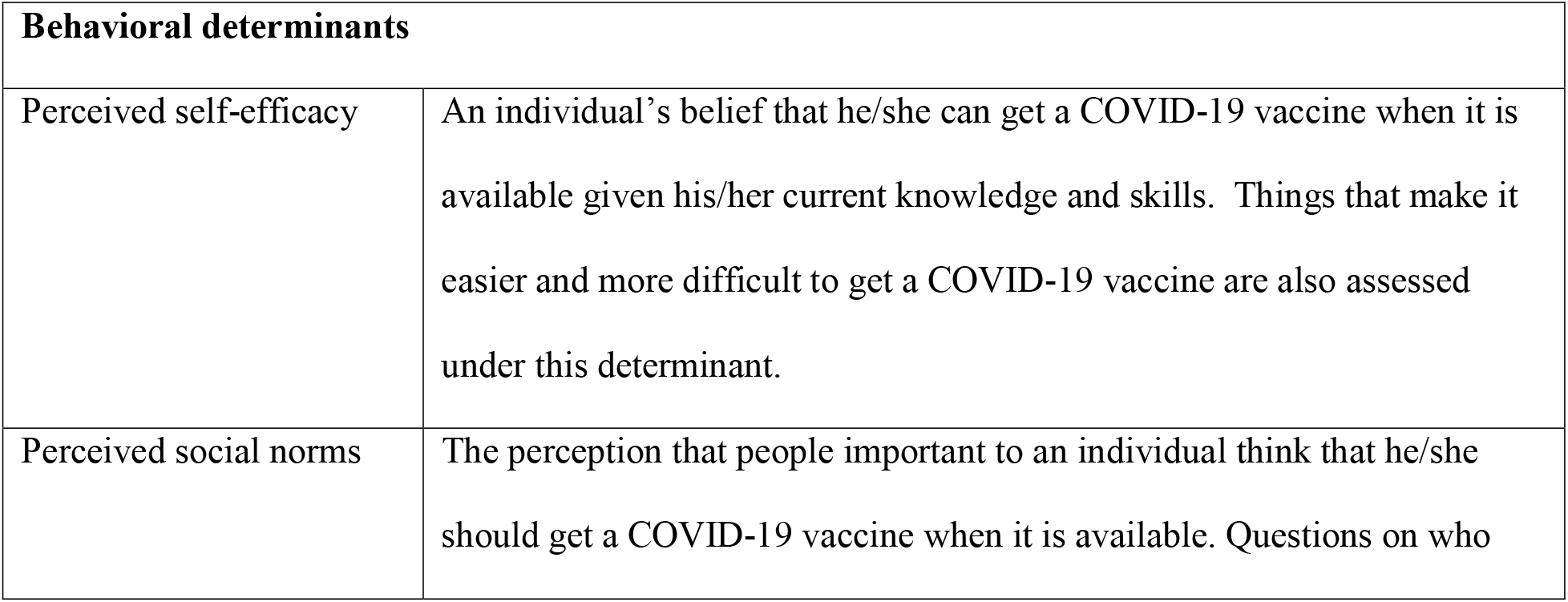
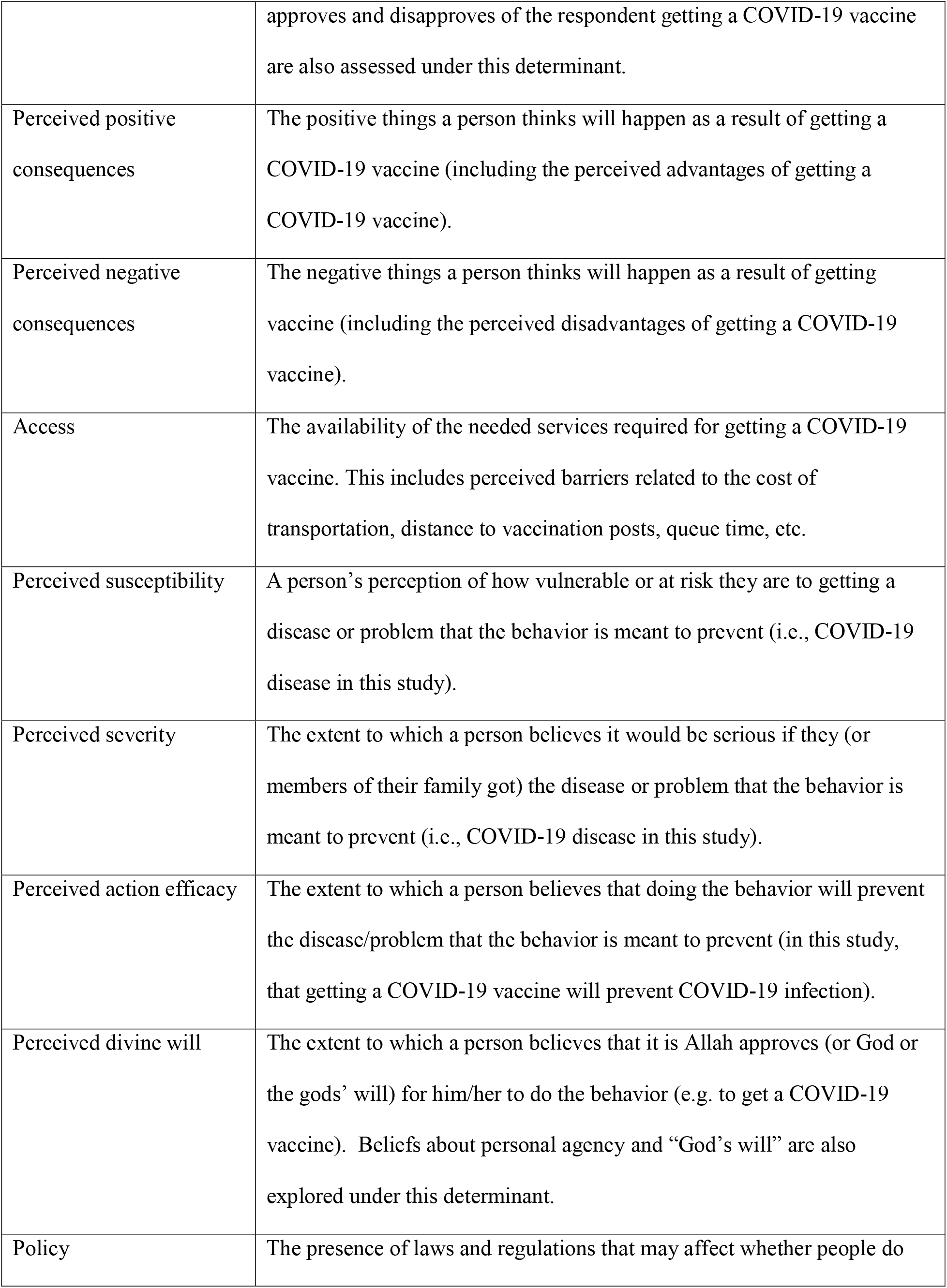
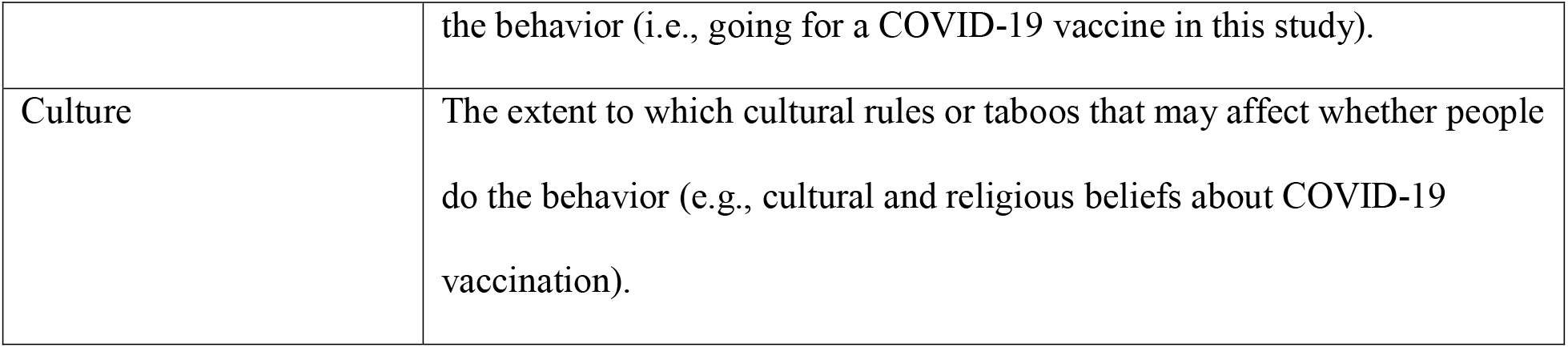

The BA questionnaire is divided into three parts. The first part includes a set of screening questions to identify whether the participant as either an ‘Acceptor’ or a ‘Non-acceptor’. In the screening section of the questionnaire, self-reported intention questions were used. (Please see Supplementary file 1.) In the second part of the questionnaire, information on the respondents’ age, gender, level of education and profession are obtained. In order to assess 11 of the usual 12 determinants assessed with BA (see Box 1), the third section consists of a set of open- and closed-ended questions (37,44). Based on input and a literature review from NGO experts who worked on increasing vaccine acceptance in the Ebola Vaccine Deployment and Compliance Project, several questions were also added to the standard BA questionnaire to explore respondent’s beliefs on the safety and effectiveness of COVID-19 vaccines, trust in vaccine information provided by both community and religious leaders, exposure to misinformation, and level of education. Based on local social media listening, one question was added on perceptions around herd immunity. The questionnaire was developed in English and then was translated into Bengali and back-translated into English to check for accuracy.

### Sampling

We interviewed adult men and women for this BA study and selected them randomly through a convenience sampling strategy from five different areas of Dhaka – the capital city of Bangladesh. The Barrier Analysis approach recommends a minimum sample size of 45 Doers (Acceptors) and 45 Non-doers (Non-acceptors) in order to detect statistically-significant Odds Ratios of 3.0 or higher, an alpha error of 5%, and a power of 80% (37). The data collection team approached adult men and women until they reached 90 respondents (45 Acceptors and 45 Non-acceptors).

### Data collection, management and analysis

We collected data between 9 to 13 January 2021 through individual interviews with responses recorded on paper-based questionnaires by two teams composed of one female and one male member. Male enumerators interviewed male respondents, and female enumerators interviewed female respondents. A research supervisor assured the quality of data. Following data collection, the data collection team and the lead author coded the open-ended responses thematically, using both an inductive and deductive coding process. At the end of this process, the team quantified the responses in each category for Acceptors and Non-acceptors separately. These categories and the number of responses registered for each were then entered into a standardized BA tabulation sheet that revealed whether differences in the proportion of Acceptors and Non-acceptors providing each response were statistically significant and should be addressed through the behavior change strategy. For each question and category of responses, the BA tabulation calculates the percentage of responses for both Acceptors and Non-Acceptors; the Odds Ratio, the Standard Error, and its confidence interval; the Estimated Relative Risk (ERR); and p-values (see supplementary file 2). This allows identify those differences between Acceptors and Non-acceptors that are statistically significant (at p<0.01) and to see the strength of the associations between each response and the behavior (based on the ERR).

### Ethical considerations

We performed all procedures in studies in accordance with the ethical standards of the institutional and/or national research committee and with the 1964 Helsinki Declaration and its later amendments or comparable ethical standards. The study protocol was approved by the institutional Ethics Committee of the Chattogram Veterinary and Animal Sciences University, Bangladesh (permit ref. no. CVASU/Dir (R and E) EC/2020/169). We informed respondents about the study objectives, and obtained their written consent before conducted interview. The data collection activities have performed following the COVID-19 safety protocols in Bangladesh that enacted by Directorate General of Health Services in Bangladesh (50).

## 3. Results

The statistically significant differences in responses and beliefs were found between Acceptors and Non-acceptors of COVID-19 vaccine are shown below. The categories of determinants are organized from higher to lower estimated relative risk ratios (ERR).

### 3.1 Perceived Social Norms

Some of the strongest predictors of vaccine acceptance in this population are beliefs around both injunctive and descriptive social norms, who the respondent thinks approve or disapproves of COVID-19 vaccination, and the proportion of people that they think will go for a COVID-19 vaccine when it is available. The results are shown in Table 1. Specifically, Acceptors were 3.2 times more likely to say they would be “very likely” to get a COVID-19 vaccine if a doctor or nurse approved (p<0.001), while Non-acceptors were 2.6 more likely to say it would be “not likely” that they would get a vaccine if a doctor or nurse recommended it (p<0.001). Acceptors were twice as likely to say that “most people” they know will get a vaccine (p<0.001), and 1.3 times more likely to say that “most close family and friends” will get a vaccine (p=0.003), while conversely, Non-acceptors were 3.5 times more likely to say that “very few people” they knew would get a vaccine (p<0.001) and 1.3 times more likely to say that “most of their close family and friends” would not get a COVID-19 vaccine (p=0.003). In terms of respondents’ impressions concerning who disapproves of their getting a COVID-19 vaccine, Acceptors were 1.7 times more likely (than Non-acceptors) to say that “no one” would disapprove (p<0.001). Non-acceptors were 1.5 times more likely to say that “my mother” (p<0.004), 1.4 times more likely to say “elderly people” (p=0.009), and 1.7 times more likely to say “people who will not get the vaccine” (p=0.006) would disapprove of their getting a COVID-19 vaccine. In addition, Acceptors were and more likely to say their mother would approve of their getting a COVID-19 vaccine (p<0.001). With regards to community and religious leaders’ influence on the decision to get a COVID-19 vaccine, Acceptors were 1.3 times more likely to say that “most community and religious leaders” would want them to get a vaccine (p=0.007), while Non-acceptors were 1.5 times more likely to say that most community leaders and religious leaders would not want them to get a COVID-19 vaccine (p=0.002).

**Table 1:**
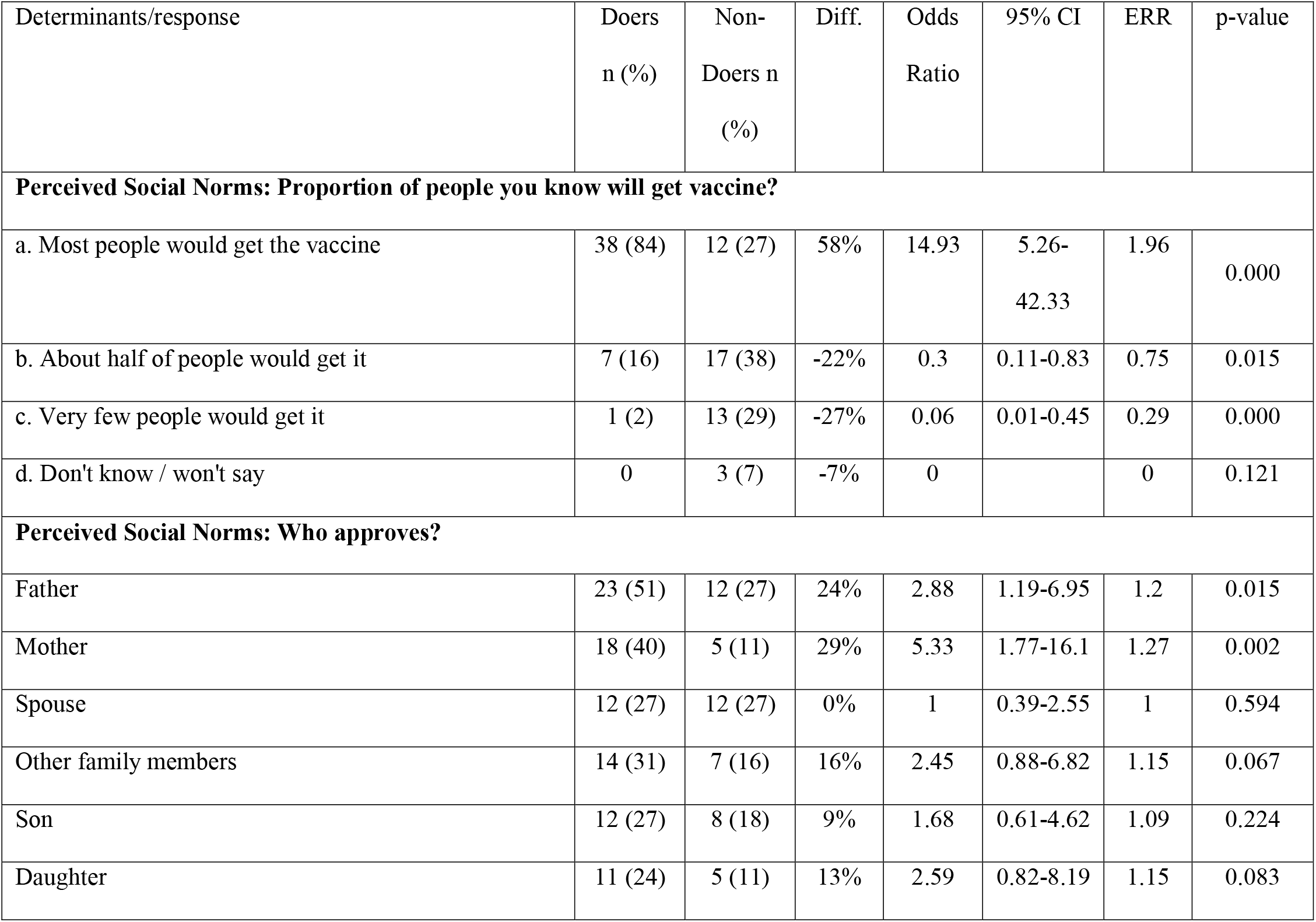
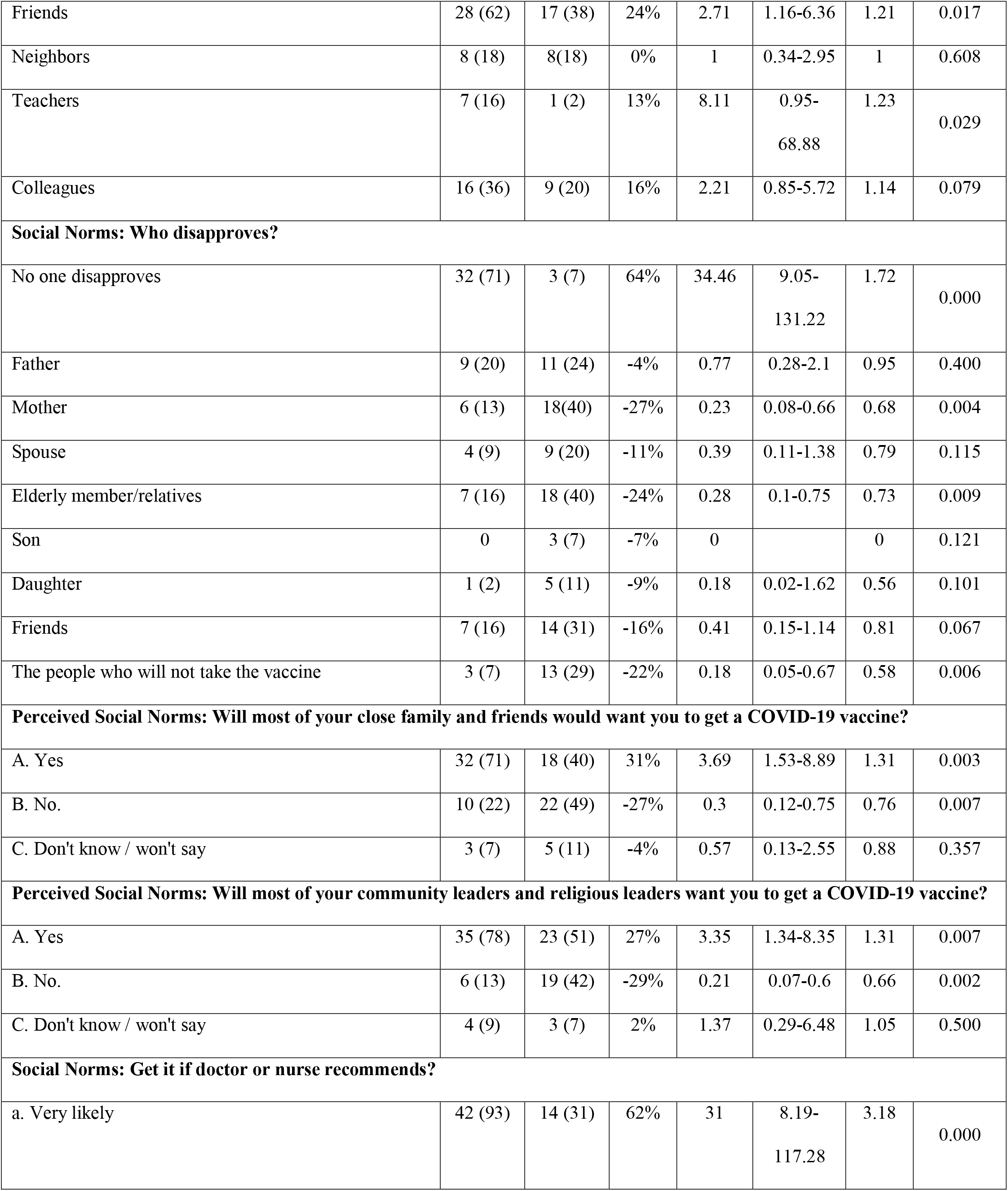
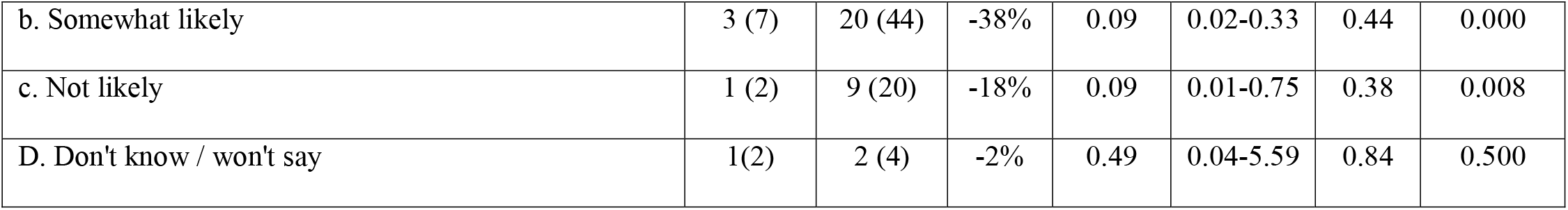
Perceived Social Norms.

### 3.2 Perceived Safety of COVID-19 Vaccines

As in many places in the world, concerns about safety of COVID-19 vaccines are affecting intention to accept vaccination in this population. When asked how safe the COVID-19 vaccines are, Non-acceptors were 1.8 times more likely to say that COVID-19 vaccines are “not safe at all” (p<0.001) while Acceptors were 1.4 times more likely to say that COVID-19 vaccines are “mostly safe” (p<0.001) (Table 2).

**Table 2:**
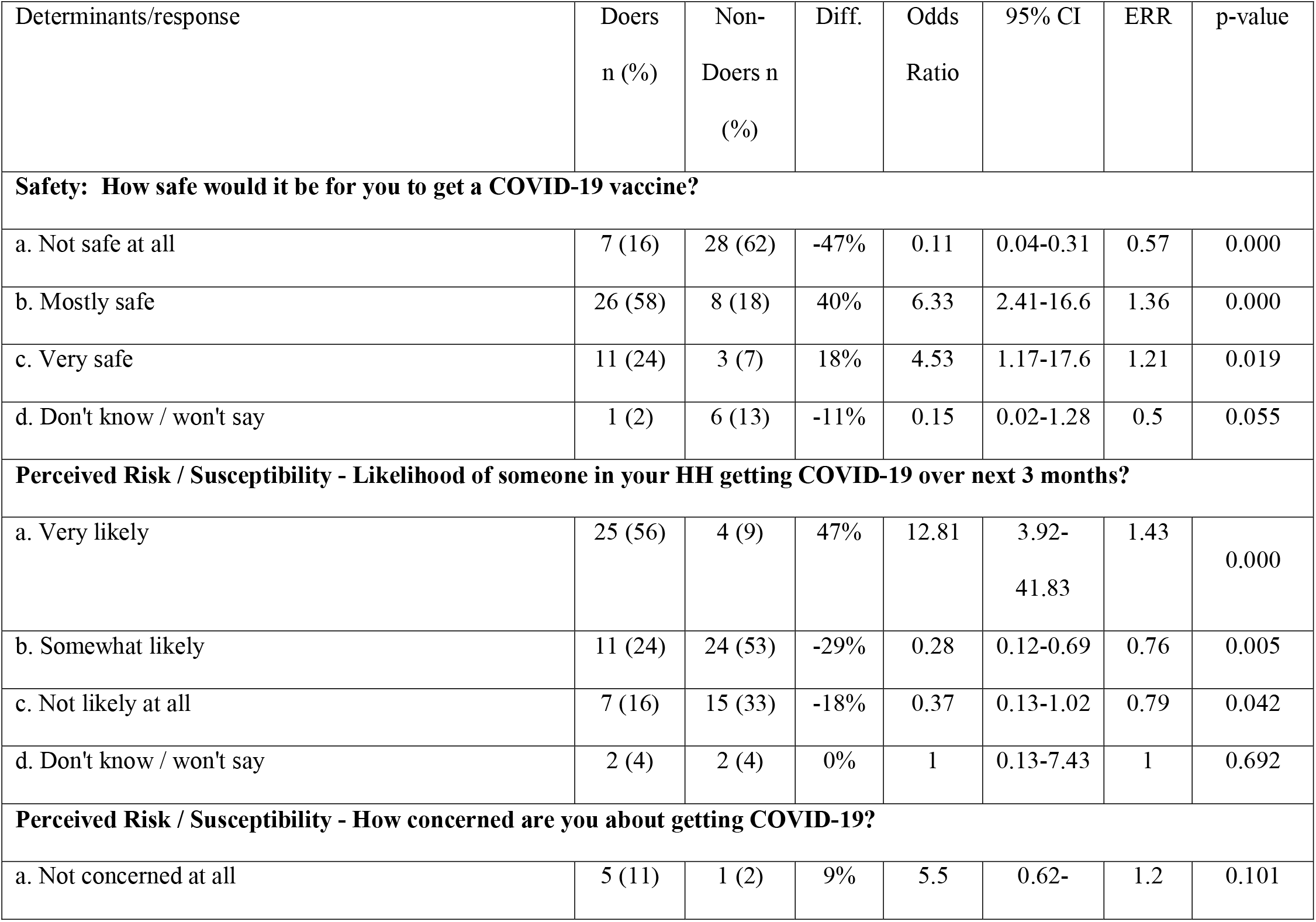
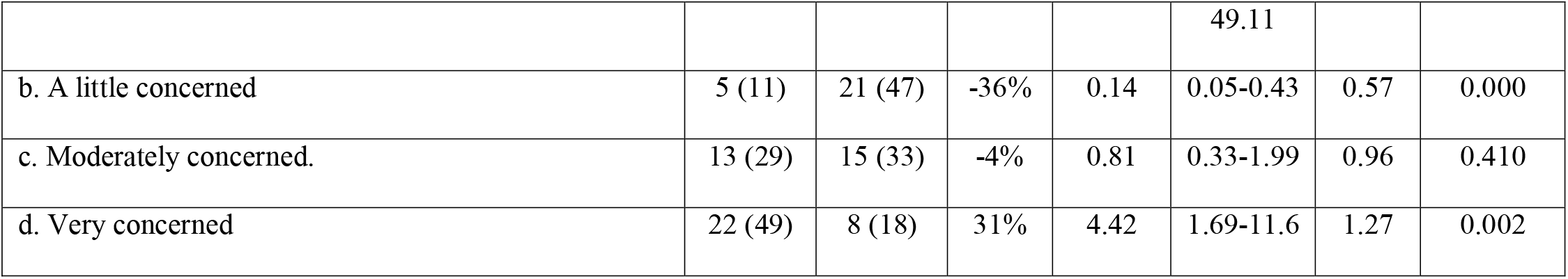
Perceived safety and risk.

### 3.3 Perceived Risk / Susceptibility (to COVID-19)

Perceived risk of getting COVID-19 – and the level of emotional concern about getting COVID-19 – also appears to be driving intended vaccine acceptance in Dhaka. Acceptors were 1.4 times more likely to say they it was “very likely” that someone in their household would get COVID-19 over the next 3 months (p<0.001) while Non-acceptors were 1.3 times more likely to say that was only “somewhat likely” (p=0.005). When respondents were asked how concerned they were about getting COVID-19 (or someone in their household getting COVID-19), Acceptors were 1.3 times more likely to say that they were “very concerned” (p=0.002) while Non-acceptors were 1.7 times more likely to say that they were only “a little concerned” (p<0.001). Additionally, Non-acceptors were 1.7 times more likely to say that “very few people” have had COVID-19 in their communities (p<0.001) (Table 2).

### 3.4 Perceived Self-efficacy

The respondents were asked two open-ended questions to understand what would make it easier or more difficult to get a COVID-19 vaccine. From the results (Table 3), factors related to *how* and *where* the vaccine would be given affects intention to vaccinate, including whether or not proper COVID-19 social distancing and prevention measures are maintained during vaccination.

**Table 3:** Perceived self-efficacy.

| Determinants/response | Doers<br>n (%) | Non-Doers n<br>(%) | Diff. | Odds<br>Ratio | 95% CI | ERR | p-value |
| --- | --- | --- | --- | --- | --- | --- | --- |
| **Self-Efficacy: What would make it easier?** |  |  |  |  |  |  |  |
| If vaccine is provided by government health centers or hospitals | 23 (51) | 5 (11) | 40% | 8.36 | 2.79-25.08 | 1.36 | 0.000 |
| School-based vaccination centers | 21 (47) | 8 (18) | 29% | 4.05 | 1.55-10.60 | 1.25 | 0.003 |
| If proper health and safety (COVID-19) protocols are maintained while giving vaccine | 20 (44) | 8 (18) | 27% | 3.7 | 1.41-9.70 | 1.23 | 0.006 |
| If the vaccines are given at home | 15 (33) | 27 (60) | -27% | 0.33 | 0.14-0.79 | 0.8 | 0.010 |
| If the vaccines are provided by establishing kiosks | 18 (40) | 4 (9) | 31% | 6.83 | 2.08-22.40 | 1.29 | 0.001 |
| When the vaccine is truly effective | 12 (27) | 15 (33) | -7% | 0.73 | 0.29-1.80 | 0.94 | 0.323 |
| If the vaccines are provided in every house | 15 (33) | 27 (60) | -27% | 0.33 | 0.14-0.79 | 0.8 | 0.010 |
| If vaccines are given by experienced health workers | 14 (31) | 9 (20) | 11% | 1.81 | 0.69-4.74 | 1.11 | 0.167 |
| Community/Para-based vaccination campaign | 10 (22) | 5 (11) | 11% | 2.29 | 0.71-7.33 | 1.13 | 0.129 |
| Awareness raising on the positive aspects through social media, TV, mike | 9 (20) | 16 (36) | -16% | 0.45 | 0.17-1.17 | 0.84 | 0.079 |
| When the vaccines do not have any side effects | 4 (9) | 16 (36) | -27% | 0.18 | 0.05-0.58 | 0.6 | 0.002 |
| Mosque-based vaccination program | 8 (18) | 5 (11) | 7% | 1.73 | 0.52-5.76 | 1.09 | 0.275 |
| Prior notice on the schedule and maintaining that schedule | 16 (36) | 7 (20) | 20% | 3 | 1.09-8.23 | 1.19 | 0.026 |
| Equal opportunity | 8 (18) | 6 (13) | 4% | 1.41 | 0.44-4.44 | 1.06 | 0.386 |
| Vaccine distribution by NGO | 8 (18) | 4 (9) | 9% | 2.22 | 0.62-7.97 | 1.13 | 0.176 |
| Enough human resources | 12 (27) | 10 (22) | 4% | 1.27 | 0.49-3.34 | 1.04 | 0.403 |
| Having vaccine at the workplace | 3 (7) | 5 (11) | -4% | 0.57 | 0.13-2.55 | 0.88 | 0.357 |
| **Self-Efficacy: What would make it Difficult?** |  |  |  |  |  |  |  |
| If the vaccinators are not skilled | 16 (36) | 6 (13) | 22% | 3.59 | 1.25-103 | 1.21 | 0.013 |
| If there are no health measures in the vaccination center due to overcrowd (Risk of getting infected with COVID-19 while vaccinating) | 18 (40) | 4 (9) | 31% | 6.83 | 2.08-22.4 | 1.29 | 0.001 |
| If the vaccine is not effective | 13 (29) | 22 (49) | -20% | 0.42 | 0.18-101 | 0.84 | 0.041 |
| If the vaccinator does not follow proper COVID-19 preventive measures | 19 (42) | 7 (16) | 27% | 3.97 | 1.46-10.8 | 1.24 | 0.005 |
| If there is corruption, syndicate and misuse of power while distribution | 13 (29) | 19 (42) | -13% | 0.56 | 0.23-1.33 | 0.89 | 0.135 |
| If the vaccines are provided by organizations other than government hospitals or health centers | 6 (13) | 9 (20) | -7% | 0.62 | 0.2-1.9 | 0.9 | 0.286 |
| If it requires more time to get vaccinated | 4 (9) | 9 (20) | -11% | 0.39 | 0.11-1.38 | 0.79 | 0.115 |
| If the vaccination center is far from home | 12 (27) | 8 (18) | 9% | 1.68 | 0.61-4.62 | 1.09 | 0.224 |
| When the vaccine has severe side-effects | 3 (7) | 22 (49) | -42% | 0.07 | 0.02-0.28 | 0.41 | 0.000 |
| If there is bribery | 2 (4) | 12 (27) | -22% | 0.13 | 0.03-0.61 | 0.49 | 0.004 |
| If political leaders influence to get vaccine to their favorite people | 5 (11) | 8 (18) | -7% | 0.58 | 0.17-1.93 | 0.88 | 0.275 |

Concerning what made it **easier**, Acceptors were 1.4 times more likely to say “if vaccination is provided by government health care facilities” (p=0.000), 1.2 times more likely to say “school-based vaccination” (p=0.003), 1.3 times more likely to say “if the vaccines are provided by establishing kiosks” (p=0.001) and 1.2 times more likely to say “if proper health & safety (COVID-19) protocols are maintained while giving the vaccine” (p=0.006) all would make it easier to get a COVID-19 vaccine. Conversely – and interestingly – Non-acceptors were 1.2 times more likely to say “when vaccines are given at home” (p=0.010), and 1.7 times more likely to say “when the vaccine has no side effects” (p=0.002) would make it easier for them to get a COVID-19 vaccine.

When asked what would make it more difficult to get a COVID-19 vaccine, Acceptors were 1.3 times more likely to say either “if there are no health measures in the vaccination center due to overcrowding” or “risk of getting infected with COVID-19 while vaccinating” (p=0.001), and 1.2 times more likely to say “if the vaccinator does not follow proper COVID-19 preventive measures” (p=0.005) would make it difficult to get a COVID-19 vaccine. Non-acceptors were 2.4 times more likely to say “when the vaccine has severe side-effects” (p<0.001) and 2 time more likely to say “if there is bribery” (p=0.004) would make it difficult to get a COVID-19 vaccine.

### 3.5 Perceived Positive Consequences and Perceived Negative Consequences

Respondents were also asked what the positive and negative consequences (e.g., advantages and disadvantages) would be of getting a COVID-19 vaccine. While reducing the reduced risk of infection was important, Acceptors were more likely to point out benefits related to livelihood and economic benefits and life getting back to normal. Specifically (Table 4), Acceptors were 1.3 times more likely to say that reducing the risk of COVID-19 infection (p=0.003), 1.3 times more likely to say “we can attend social and cultural activities” (p=0.003), 1.2 times more likely to say “children can start school again” (p=0.003), 1.3 times more likely to say “reduction in COVID-19 related costs” (e.g. masks, hand sanitizer, tests; p<0.001), 1.3 times more likely to say “employment and income opportunities will be increased” (p<0.001), and 1.3 times more likely to say “attending prayers in congregation” (p<0.002) were all reported advantages of getting a COVID-19 vaccine.

**Table 4:** Perceived positive and negative consequences.

| Determinants/response | Doers<br>n (%) | Non-Doers n<br>(%) | Diff. | Odds<br>Ratio | 95% CI | ERR | p-value |
| --- | --- | --- | --- | --- | --- | --- | --- |
| **Perceived Positive Consequences (Advantages)** |  |  |  |  |  |  |  |
| Reduce the risk of COVID infection | 30 (67) | 16 (36) | 31% | 3.63 | 1.52-8.65 | 1.29 | 0.003 |
| Our health will be protected | 18 (40) | 9 (20) | 20% | 2.67 | 1.04-6.85 | 1.17 | 0.032 |
| Can move easily | 14 (31) | 15 (33) | -2% | 0.9 | 0.37-2.19 | 0.98 | 0.500 |
| Will get fearless life | 17 (38) | 14 (31) | 7% | 1.34 | 0.56-3.22 | 1.06 | 0.329 |
| We can attend social and cultural activities | 14 (31) | 3 (7) | 24% | 6.32 | 1.67-23.9 | 1.25 | 0.003 |
| Elderly and children can easily move here and there | 8 (18) | 3 (7) | 11% | 3.03 | 0.75-12.3 | 1.16 | 0.098 |
| Children can start their school again | 17 (38) | 5 (11) | 27% | 4.86 | 1.6-14.7 | 1.25 | 0.003 |
| All economic activities will get back to normal and increase | 19 (42) | 9 (20) | 22% | 2.92 | 1.14-7.48 | 1.19 | 0.020 |
| Reduction in COVID-19-related costs (mask, hand sanitizer, detergents, primary medicine, COVID-19 test etc). | 16 (36) | 3 (7) | 29% | 7.72 | 2.06-28.9 | 1.28 | 0.001 |
| Anxiety free life | 9 (20) | 2 (4) | 16% | 5.38 | 1.09-26.5 | 1.22 | 0.025 |
| New Employment/income source will be increased | 17 (38) | 3 (7) | 31% | 8.5 | 2.28-31.7 | 1.3 | 0.000 |
| Use of public transportation | 5 (11) | 2 (4) | 7% | 2.69 | 0.49-14.6 | 1.14 | 0.217 |
| Family members will be safe | 12 (27) | 8 (18) | 9% | 1.68 | 0.61-4.62 | 1.09 | 0.224 |
| Attend prayers in congregation | 16 (36) | 4 (9) | 27% | 5.66 | 1.71-18.7 | 1.26 | 0.002 |
| Won't need to follow health measures (social distance, wear mask and frequent handwashing) | 23 (51) | 12 (27) | 24% | 2.88 | 1.19-6.95 | 1.2 | 0.015 |
| No positive outcome | 0 | 5 (11) |  |  |  |  |  |
| **Perceived Negative Consequences (Disadvantages)** |  |  |  |  |  |  |  |
| Weakness | 23 (51) | 9 (20) | 31% | 4.18 | 1.64-10.7 | 1.27 | 0.002 |
| Body ache | 15 (33) | 5 (11) | 22% | 4 | 1.31-12.2 | 1.22 | 0.010 |
| Women may get beard and moustache | 4 (9) | 13 (29) | -20% | 0.24 | 0.07-0.81 | 0.67 | 0.015 |
| Fever | 18 (40) | 12 (27) | 13% | 1.83 | 0.75-4.46 | 1.11 | 0.132 |
| Itching and skin problem | 17 (38) | 3 (7) | 31% | 8.5 | 2.28-31.7 | 1.3 | 0.000 |
| Life threatening side effects | 7 (16) | 24 (53) | -38% | 0.16 | 0.06-0.44 | 0.63 | 0.000 |
| Unknown diseases | 6 (13) | 18 (40) | -27% | 0.23 | 0.08-0.66 | 0.68 | 0.004 |
| Different chronic diseases (Cancer, Kidney, diabetes) | 9 (20) | 19 (42) | -22% | 0.34 | 0.13-0.88 | 0.78 | 0.020 |
| Infertility | 3 (7) | 15 (33) | -27% | 0.14 | 0.04-0.54 | 0.54 | 0.001 |
| Convulsion | 4 (9) | 11 (24) | -16% | 0.3 | 0.09-1.03 | 0.73 | 0.044 |
| Hormonal imbalance | 5 (11) | 7 (16) | -4% | 0.68 | 0.2-2.32 | 0.92 | 0.379 |
| Vomiting and dizziness | 8 (18) | 4 (9) | 9% | 2.22 | 0.62-7.97 | 1.13 | 0.176 |
| People would get reluctant on following health measures | 9 (20) | 3 (7) | 13% | 3.5 | 0.88-13.92 | 1.18 | 0.059 |
| **Perceived Social Norms: Proportion of people you know will get vaccine?** |  |  |  |  |  |  |  |
| a. Most people would get the vaccine | 38 (84) | 12 (27) | 58% | 14.93 | 5.26-42.33 | 1.96 | 0.000 |
| b. About half of people would get it | 7 (16) | 17 (38) | -22% | 0.3 | 0.11-0.83 | 0.75 | 0.015 |
| c. Very few people would get it | 1 (2) | 13 (29) | -27% | 0.06 | 0.01-0.45 | 0.29 | 0.000 |
| d. Don't know / won't say | 0 | 3 (7) | -7% | 0 |  | 0 | 0.121 |

**Table 5:** Perceived action efficacy and trust in COVID-19 vaccine.

| Determinants/response | Doers<br>n (%) | Non-Doers n<br>(%) | Diff. | Odds<br>Ratio | 95% CI | ERR | p-value |
| --- | --- | --- | --- | --- | --- | --- | --- |
| **Action Efficacy - Likelihood of getting COVID-19 after getting COVID-19 vaccine?** |  |  |  |  |  |  |  |
| a. Very likely | 11 (24) | 3 (7) | 18% | 4.53 | 1.17-17.6 | 1.21 | 0.019 |
| b. Somewhat likely | 22 (49) | 6 (13) | 36% | 6.22 | 2.2-17.6 | 1.31 | 0.000 |
| c. Not likely at all | 2 (4) | 19 (42) | -38% | 0.06 | 0.01-0.3 | 0.35 | 0.000 |
| d. Don't know / won't say | 10 (22) | 17 (38) | -16% | 0.47 | 0.19-1.19 | 0.85 | 0.083 |
| **Trust (in information) - How much would you trust a COVID-19 vaccine?** |  |  |  |  |  |  |  |
| a. Not trust it at all | 2 (4) | 14 (31) | -27% | 0.1 | 0.02-0.49 | 0.44 | 0.001 |
| b. Trust it a little | 6 (13) | 23 (51) | -38% | 0.15 | 0.05-0.42 | 0.6 | 0.000 |
| c. Trust it a moderate amount | 19 (42) | 6 (13) | 29% | 4.75 | 1.67-13.5 | 1.26 | 0.002 |
| d. Trust it a lot | 18 (40) | 2 (4) | 36% | 14.33 | 3.08-66.7 | 1.34 | 0.000 |

When asked about the negative consequences (disadvantages), Acceptors were 1.3 times more likely to mention mild side effects of vaccination such as “weakness” (p<0.002) and 1.3 times more likely to say “itching and skin problems” (p<0.001) were disadvantages of getting a COVID-19 vaccine. Meanwhile, Non-acceptors were 1.6 times more likely (than Acceptors) to mention life-threatening side effects (p<0.001), 1.5 times more likely to say “unknown/new diseases” (p<0.004), and 1.9 times more likely to say “infertility” (p<0.001) were all disadvantages of getting a COVID-19 vaccine.

### 3.6 Perceived Action Efficacy

Counter-intuitively, Acceptors were 1.3 times more likely to say they were “somewhat likely” to get COVID-19 once they were vaccinated against it (p<0.001) while Non-acceptors were 2.8 times more likely to say that they were “not likely at all” to get COVID-19 once one was vaccinated against it (p<0.001) (Table **5**). Focus group discussions may be used at a later point in time to explore this finding. Related to perceived action efficacy, respondents were asked if they agreed or disagreed with the statement “Most people will eventually get infected with COVID-19, so getting a COVID-19 vaccine is unnecessary.” Non-acceptors were 1.7 times more likely to say that they “agree a little” or “agree a lot” with the statement (p<0.001) while Acceptors were 1.5 times more likely to “disagree a lot” with the statement (p<0.001).

### 3.7 Trust in COVID-19 vaccines

As expected, trust in COVID-19 vaccines is a key driver of intended vaccine acceptance in Dhaka. Acceptors were twice as likely to say that they trust the COVID-19 vaccines “a lot” or a “moderate amount” (p<0.001). Conversely, Non-acceptors were 1.7 times more likely to say that they “trust them a little” (p<0.001) and 2.3 times more likely to say that they “do not trust [COVID-19 vaccines] at all” (p=0.001) (Table 6).

**Table 6:** Perceived severity and perceived access.

| Determinants/response | Doers<br>n (%) | Non-Doers n<br>(%) | Diff. | Odds<br>Ratio | 95% CI | ERR | p-value |
| --- | --- | --- | --- | --- | --- | --- | --- |
| **Severity - How serious if someone in your HH got COVID-19?** |  |  |  |  |  |  |  |
| a. Very serious | 27 (60) | 12 (27) | 33% | 4.13 | 1.69-10.1 | 1.3 | 0.001 |
| b. Somewhat serious | 10 (22) | 11 (24) | -2% | 0.88 | 0.33-2.35 | 0.98 | 0.500 |
| c. Not serious at all | 8 (18) | 16 (36) | -18% | 0.39 | 0.15-1.04 | 0.81 | 0.047 |
| d. Don't know / won't say | 0 | 6 (13) | -13% | 0 |  | 0 | 0.013 |

| **Access - How difficult for you to get to the clinic where vaccines are normally offered?** |  |  |  |  |  |  |  |
| --- | --- | --- | --- | --- | --- | --- | --- |
| A. Very difficult | 8 (18) | 28 (62) | -44% | 0.13 | 0.05-0.35 | 0.61 | 0.000 |
| B. Somewhat difficult | 20 (44) | 5 (11) | 33% | 6.4 | 2.13-19.23 | 1.3 | 0.000 |
| C. Not difficult at all | 17 (38) | 12 (27) | 11% | 1.67 | 0.68-4.08 | 1.1 | 0.184 |
| **Access - Expected queue time** |  |  |  |  |  |  |  |
| A. 0-30 mins | 7 (16) | 7 (16) | 0% | 1 | 0.32-3.13 | 1 | 0.614 |
| B. 31-60 mins | 1 (2) | 4 (9) | -7% | 0.23 | 0.02-2.17 | 0.63 | 0.180 |
| C. 1 hr - 1.5 hrs | 3 (7) | 6 (13) | -7% | 0.46 | 0.11-1.98 | 0.83 | 0.242 |
| D. 1.5 - 2.0 hrs | 12 (27) | 3 (7) | 20% | 5.09 | 1.33-19.54 | 1.23 | 0.011 |
| E. 2-3 hrs | 22 (49) | 25 (56) | -7% | 0.77 | 0.33-1.75 | 0.95 | 0.337 |

### 3.8 Perceived Severity (of COVID-19)

The **perceived severity** of COVID-19 was also highly correlated with intended vaccine acceptance. Respondents were asked how serious it would be if they or someone else in their household got COVID-19. Acceptors were 1.3 times more likely (than Non-acceptors) to say that it would be “very serious” (p=0.001) (Table 6).

### 3.9 (Perceived) Access

Perceived difficultly in reaching clinics that normally provide vaccines is associated with intended COVID-19 vaccine acceptance in Dhaka. Non-acceptors were 1.6 times more likely to say that it was “very difficult” to get to the facility that normally provides vaccines (p=0.000), while Acceptors were 1.3 times more likely to say it was “somewhat difficult” to get to that facility (p=0.000) (Table 6).

### 3.10 Perceived Divine Will

Personal agency and religious beliefs often come into play with vaccine acceptance. In this study (Table 7), we asked for respondents’ degree of agreement or disagreement with the statement: “Whether I get COVID-19 or not is purely a matter of God’s will or chance -- The actions I take will have little bearing on whether or not I get COVID-19.” Personal agency was correlated with intended vaccine acceptance: Acceptors were 1.2 times more likely to say that they “disagree a lot” (p=0.005) that “whether I get COVID-19 or not is purely a matter of God’s will or chance” (p<0.001). Respondents were also asked whether they believed that Allah (or God or the gods) approves or disapproves of people getting COVID-19 vaccines. While 80% of Acceptors and 78% of Non-acceptors said that a deity approves, there were no statistically significant differences between Acceptors and Non-acceptors for this question.

**Table 7:** Perceived divine will and culture/rumors.

| Determinants/response | Doers<br>n (%) | Non-Doers n<br>(%) | Diff. | Odds<br>Ratio | 95% CI | ERR | p-value |
| --- | --- | --- | --- | --- | --- | --- | --- |
| **Divine Will - Does God/Allah/the gods approve or disapprove of people getting COVID-19 vaccine?** |  |  |  |  |  |  |  |
| a. Believe God/Allah/gods approve | 36<br>(80) | 35 (78) | 2% | 1.14 | 0.41-3.15 | 1.03 | 0.500 |
| b. Believe God/Allah/gods does not approve | 1 (2) | 0 | 2% |  |  | 1.24 | 0.500 |
| c. Believe God/Allah/gods does not approve or disapprove | 0 | 3 (7) | -7% | 0 |  | 0 | 0.121 |
| d. Don't know / won't say | 8 (18) | 7 (16) | 2% | 1.17 | 0.39-3.56 | 1.03 | 0.500 |
| **Divine Will - Agree/disagree with "whether I get COVID-19 or not is purely a matter of God's will or chance.** |  |  |  |  |  |  |  |
| a. Agree a little | 10<br>(22) | 15 (33) | -11% | 0.57 | 0.22-1.46 | 0.89 | 0.173 |
| b. Agree a lot | 2 (4) | 4 (9) | -4% | 0.48 | 0.08-2.74 | 0.83 | 0.338 |
| c. Disagree a little | 7 (16) | 13 (29) | -13% | 0.45 | 0.16-0.83 | 0.83 | 0.102 |
| d. Disagree a lot | 26<br>(58) | 13 (29) | 29% | 3.37 | 1.4-8.08 | 1.25 | 0.005 |
| **Culture - Any cultural or religious reasons you would not get COVID-19 vaccine?** |  |  |  |  |  |  |  |
| a. Yes | 11 (24) | 25 (56) | -31% | 0.26 | 0.11-0.64 | 0.74 | 0.002 |
| b. No | 30 (67) | 18 (40) | 27% | 3 | 1.27-7.09 | 1.25 | 0.010 |
| c. Don't know / won't say | 1 (2) | 2 (4) | -2% | 0.49 | 0.04-5.59 | 0.84 | 0.500 |
| **(If yes to Culture) What reasons** |  |  |  |  |  |  |  |
| Use of pork fat while making vaccine- Islam does not allow this. | 5 (11) | 19 (42) | -31% | 0.17 | 0.06-0.51 | 0.61 | 0.001 |
| Use of haram ingredients in the vaccine | 8 (18) | 21 (47) | -29% | 0.25 | 0.09-0.65 | 0.71 | 0.003 |
| **Have you seen or heard of anything that would stop you or others from seeking to get the COVID-19 vaccines?** |  |  |  |  |  |  |  |
| a. Yes | 33 (73) | 28 (62) | 11% | 1.67 | 0.68-4.08 | 1.11 | 0.184 |
| b. No | 12 (27) | 17 (38) | -11% | 0.6 | 0.24-1.46 | 0.9 | 0.184 |
| **If Yes to rumors) What would stop you or others from seeking the vaccine?** |  |  |  |  |  |  |  |
| Producers' hide and seek activities related to vaccine accuracy in the clinical test | 16 (36) | 4 (9) | 27% | 5.66 | 1.71-18.7 | 1.26 | 0.002 |
| The virus will infect us even if we get vaccinated. | 3 (7) | 9 (20) | -13% | 0.29 | 0.07-1.14 | 0.7 | 0.059 |
| Some Islamic leader may prohibit getting the vaccine | 4 (9) | 7 (16) | -7% | 0.53 | 0.14-1.95 | 0.86 | 0.261 |
| It is not possible to produce a vaccine within a year. So, this vaccine may not work | 3 (7) | 12 (27) | -20% | 0.2 | 0.05-0.75 | 0.61 | 0.011 |
| Vaccine may not work against other strains [variants] | 23 (51) | 12 (27) | 24% | 2.88 | 1.19-6.95 | 1.2 | 0.015 |
| Concern about the efficacy as many volunteers got sick/died as a result of getting the vaccines | 16 (36) | 26 (58) | -22% | 0.4 | 0.17-0.94 | 0.84 | 0.028 |
| There is no proper cold chain to preserve the vaccine, so vaccine may loss its efficacy | 7 (16) | 2 (4) | 11% | 3.96 | 0.78-20.2 | 1.19 | 0.079 |

### 3.11 Rumors/ Culture

Respondents were asked if there were any cultural or religious reasons that they would not get a COVID-19 vaccine (results are in Table 7). Acceptors were 1.2 times more likely to say that there were no cultural or religious reasons they would not get a COVID-19 vaccine (p=0.01), while Non-acceptors were 1.3 more likely to say that there were reasons (p=0.002). When asked what those reasons were, Non-Acceptors were 1.6 more likely to say that they had heard that ‘the vaccines were made with pork fat which is not allowed (*haram*) by Islam’ (p=0.001) and 1.4 times more likely to say that ‘vaccines were made with haram ingredients’ (p=0.003). Respondents were also asked if they had seen or heard of anything that would stop them or others from seeking to get a COVID-19 vaccine. If they said yes, they were asked a follow-up question on what would stop them or others from seeking the vaccine. Regarding that question, Acceptors were 1.3 more likely to say that producers’ “hide and seek activities” related to vaccine accuracy in the clinical testing would stop them or their peers from getting a COVID-19 vaccine (p<0.002).

## 4. Discussion

This Barrier Analysis study on intended acceptance of COVID-19 vaccines revealed important differences in responses and beliefs between Acceptors and Non-acceptors regarding behavioral determinants of vaccine acceptance in this urban setting of Bangladesh. One important finding is that even if one or two determinants or barriers are addressed, there are a multitude of important determinants and barriers that may affect vaccine acceptance, and deserve attention. Access, for instance, was among other determinants found to be important in our study in along with perceived social norms (75% of the studies) and positive/negative consequences of the behavior (56%) (51).

The largest behavioral driver in this population, based on responses to questions on the 11 determinants studied, was around perceived social norms. Perceived social norms, depending on the context, may hinder or inspire one to get a vaccine (52). In this study, Acceptors were more likely (than Non-acceptors) to say that most people they know and most of their close family and friends will get the COVID-19 vaccine, that no one would disapprove of their getting a vaccine, and that they would be very likely to get a vaccine if a doctor or nurse recommended it. Similarly, one systematic review showed health care professionals are influential in promoting vaccinations (53). Acceptors were also more likely to say that most of their community and religious leaders will want them to get a vaccine.

Other beliefs, for example safety and trust, about the vaccines themselves were important, as well (54,55). In this study, Non-acceptors were much more likely to say that COVID-19 vaccines are “not safe at all” and to say that they only “trust them a little.” Conversely, Acceptors were more likely to say that the COVID-19 vaccines are “mostly safe” and to say that they trust them “a lot.”

In line with the Health Belief Model, beliefs about the disease itself were highly correlated with vaccine acceptance (26). Acceptors were much more likely (than Non-acceptors) to say it was “very likely” that someone in their household would get COVID-19 over the next three months and to be “very concerned” about getting COVID-19. Conversely, Non-acceptors were much more likely to say that it would be only “somewhat likely” that someone in their household would get COVID-19 and that “very few people” have had COVID-19 in their community. In alignment with what Patrick et al (56) showed regarding perceived risk as structural feature of vaccine decision, Acceptors of this study were also more likely to believe that it would be “very serious” if someone in their household got COVID-19.

Perceived behavioral control (which is influenced by things that make it difficult or easy to perform the behavior) also influence vaccine uptake (31). The barriers and enablers which were mentioned more often by Acceptors provide clues as to ways to make it easier to boost acceptance. When asked what would make it easier to get a COVID-19 vaccine, Acceptors were more likely (than Non-acceptors) to mention providing vaccination through government health facilities, schools, and kiosks, and having vaccinators maintain proper COVID-19 health & safety protocols. Responding to the question about what would make it difficult to get a COVID-19 vaccine, Non-acceptors were much more likely say “when the vaccine has severe side effects.” Personal agency also came into play: Acceptors were much more likely to say that they disagreed a lot with the statement, “whether I get COVID-19 or not is purely a matter of God’s will or chance – the actions I take will have little bearing on whether or not I get COVID-19.”

Aligned with other studies on vaccination uptake (57,58), the results of this study showed that perceived effects of vaccines are a key factor in the vaccine decision. Acceptors named several positive consequences of getting a COVID-19 vaccine more often than Non-acceptors including (1) reduced the risk of Covid-19 infection, (2) being able to attend social and cultural activities, (3) children being able to start school again, (4) reduction in COVID-19 related costs, (5) increased employment and income opportunities, and (6) being able to attend prayers in a group setting. Conversely, Non-acceptors asked about negative consequences of getting a COVID-19 vaccine were more likely to say (than Acceptors) that (1) life-threatening side effects, (2) unknown / new diseases, and (3) infertility were all disadvantages that they would expect if they were to get a COVID-19 vaccine. Surprisingly, Acceptors were 1.3 times more likely to say they were “somewhat likely” to get COVID-19 once they were vaccinated against it (p<0.001) while Non-acceptors were 2.8 times more likely to say that they were “not likely at all” to get COVID-19 if they were vaccinated.

Lastly, Non-acceptors were more likely to hold beliefs about herd immunity that could reduce acceptance, saying that they agree a little or a lot with the statement that “most people will eventually get infected with COVID-19, so getting a COVID-19 vaccine is unnecessary.”

### Implications for Behavior Change Interventions

Aligned with WHO’s Technical Advisory Group on Behavioral Insights and Sciences for Health recommendation on social and behavioral drivers on COVID-19 vaccination (33), our study identified important beliefs and responses associated with different determinants of COVID-19 vaccine acceptance among urban population in Bangladesh which could be valuable to informing contextualized behavioral intervention and engagement strategies to support COVID-19 vaccination. For example, to increase perceived positive social norms, especially for those who found to be important influencers of this behavior (e.g., medical staff and mothers), videotaping individuals giving testimonials in each neighborhood on why they plan to get the vaccine, and distributing them over media is one possible approach. Other activities can be used to make acceptance more visible (e.g., stickers on households that say, “We plan to vaccinate!” or lapel pins with the same message). To increase the perception that COVID-19 is serious (to address perceived severity), testimonials by people who have lost or almost lost family members due to COVID-19 disease could be used. To address perceived divine will, religious leaders can be assisted in creating sermon outlines on maintaining one’s health (and linking that with COVID-19 vaccines), and supported in creating radio spots to promote COVID-19 vaccines. In addition to the COVID-19 prevention, other positive consequences of immunization mentioned by Acceptors should be disseminated. While not repeating any misinformation, it will be important to provide clear information on the known minor risks of COVID-19 vaccination as a way to combat misinformation on side effects that were mentioned more often by Non-acceptors (e.g., life-threatening conditions, new diseases, infertility). Information on how vaccines are made should be disseminated to counter misinformation (e.g., that vaccines are made with pork fat or other haram ingredients). Stakeholders should also take into account the findings on things that may make vaccination easier for people, such as providing the vaccine in schools and kiosks (in addition to government health facilities) and to assure that the population understands that proper COVID-19 health and safety protocols will be maintained in places where vaccines are given.

## Limitations of the Study

This study has a number of limitations. First, given that this study was only done in a limited urban area, the results should not be generalized to the rest of Bangladesh or other countries. Second, by design (as a means to make the analysis easier for practitioners and the method replicable by more practitioners), the BA approach does not consider respondents’ socio-economic information including level of income, living conditions, or other factors which may lead to some confounding or interaction of variables. Finally, while many studies these days recognize that there is a spectrum of acceptance between those who accept, those who are undecided or hesitant, and those who refuse, for the purposes of this study and for ease of analysis, we defined vaccine acceptance in a binary way

## Conclusion

This BA study revealed a host of important behavioral determinants associated with intended COVID-19 vaccine acceptance among the study population in Dhaka, Bangladesh. Particularly, social norms, beliefs on safety and trust about the vaccines, perceived risk of getting COVID-19, perceived severity, perceived action efficacy and behavioral control. The responses were significantly varied between Acceptors and Non-acceptors which urges to address these behavioral drivers. The study also revealed some important beliefs on the positive consequences from both Acceptors and Non-acceptors, which could be used in developing behavior change messages. The results from the present study suggest that an integrated behavior change strategy needs to be incorporated into existing COVID-19 vaccination plans to increase the uptake.

## Data Availability

Data will be available based on rational request.

## Author Contribution

Conceptualization, MAK; TD, SS; methodology, MAK, TD, MMH; software, TD, MAK; validation, MAK, TD; formal analysis, MAK, TD, SS; investigation, MAK; resources, MAK, MMH, SS; data curation, TD, SS, NU, MAI; writing—original draft preparation, MAK, TD, SS; writing—review and editing, HL, TD, MAK, AI, SS, RK, MMH; supervision, TD, MMH, HL; project administration, MAK, MMH, NU, SS. All authors have read and agreed to the published version of the manuscript.

## Acknowledgments

The authors would like to acknowledge Chattogram Veterinary and Animal Sciences University for permitting to conduct this perception study. Our entire research team also grateful to all the respondents (poultry drug and feed sellers) for their kind cooperation during the interviews.

## Supporting information captions (if applicable)

